# Extracting novel antimicrobial emergence events from scientific literature and medical reports

**DOI:** 10.1101/2020.08.13.20165852

**Authors:** Emma Mendelsohn, Noam Ross, Allison M. White, Karissa Whiting, Cale Basaraba, Brooke Watson Madubuonwu, Erica Johnson, Mushtaq Dualeh, Zach Matson, Sonia Dattaray, Nchedochukwu Ezeokoli, Melanie Kirshenbaum Lieberman, Jacob Kotcher, Samantha Maher, Carlos Zambrana-Torrelio, Peter Daszak

## Abstract

Despite considerable global surveillance of antimicrobial resistance (AMR), data on the global emergence of new resistance genotypes in bacteria has not been systematically compiled. We conducted a study of English-language scientific literature (2006-2017) and disease surveillance reports (1994-2017) to identify global events of novel AMR emergence (first clinical reports of unique drug-bacteria resistance combinations). We screened 24,966 abstracts and reports, ultimately identifying 1,773 novel AMR emergence events from 294 articles. Events were reported in 66 countries, with most events in the United States (152), India (129), and China (128). The most common bacteria demonstrating new resistance were *Klebsiella pneumoniae* (352) and *Escherichia coli* (218). Resistance was most common against antibiotic drugs imipenem (89 events), ciprofloxacin (85) and ceftazidime (82). We provide an open-access database of emergence events with standardized fields for bacterial species, drugs, location, and date, and we discuss guidelines and caveats for data analysis. This database may be broadly useful for understanding rates and patterns of AMR evolution, identifying global drivers and correlates, and targeting surveillance and interventions.

## Background & Summary

Antimicrobial resistance (AMR) is a global health crisis that has compromised the effective treatment and prevention of a multitude of infections. The rise in AMR has been associated with increased mortality, longer hospitalizations, complications with medical procedures such as surgery and chemotherapy, and higher healthcare costs^1-3^. Resistance to antibiotics is a global public health issue and a particular concern in low and middle-income countries, where many high-burden diseases such as malaria, respiratory infections, and tuberculosis can no longer be treated by common antimicrobial drugs^1,4^. Combating AMR requires a multidimensional global response to optimize antimicrobial drug use, improve awareness, increase traceability and usage reporting, and promote research^1,5-7^.

AMR surveillance by researchers, hospital networks, and state governments is key to characterizing and responding to the crisis. Current global-scale datasets primarily focus on the presence or prevalence of known resistance genotypes and phenotypes in bacterial populations. In 2014, the World Health Organization (WHO) published surveillance data obtained from 129 member states on nine bacterial pathogen-antibacterial drug combinations of public health importance, finding high rates of resistance reported across the globe^3^. The ResistanceMap database [https://resistancemap.cddep.org], maintained by the Center of Disease Dynamics, Economics & Policy, provides nationally-aggregated data from 46 countries on the prevalence of resistance in 12 bacterial species against 17 classes of antibiotics^8^.

To our knowledge, however, there is no publicly available dataset that specifically identifies the spatial and temporal patterns of the emergence of novel bacterial pathogen resistance to antibiotic drugs in humans. Such data are critical to understanding macroecological patterns and drivers of AMR emergence and identifying geographic and phenotypic targets for surveillance, research, and interventions. Further, this database may support on-going agreements and programs developed by intergovernmental organizations such as the United Nations, World Health Organization, Intergovernmental Science-Policy Platform on Biodiversity and Ecosystem Services, Convention on Biological Diversity, and development agencies such as the World Bank, the Global Environment Facility, and other regional banks.

We conducted a systematic review to identify novel AMR emergence events reported in English-language scientific literature from 2006-2017 and disease surveillance reports from 1994-2017. We focused specifically on human clinical cases and included any reported resistance of a bacterium to an antimicrobial drug (i.e., we did not limit the search to a subset of bacteria or drugs). We screened 24,966 abstracts and reports, identifying 1,791 articles potentially reporting novel emergence events, and ultimately, 294 relevant studies reporting 1,773 total events. We present an open-access database of these events with cleaned and standardized fields for location of emergence, bacterial species, antimicrobial drug, emergence date, and data source. We provide usage notes on systematic biases and ambiguities in the data. In addition, we provide data on all screened and processed articles from which data were harvested.

## Methods

Development of the AMR emergence database was a multi-step process consisting of a systematic literature search, abstract and report screening, article review and coding, and data cleaning and standardization (Figure 1).

**Figure 1.**
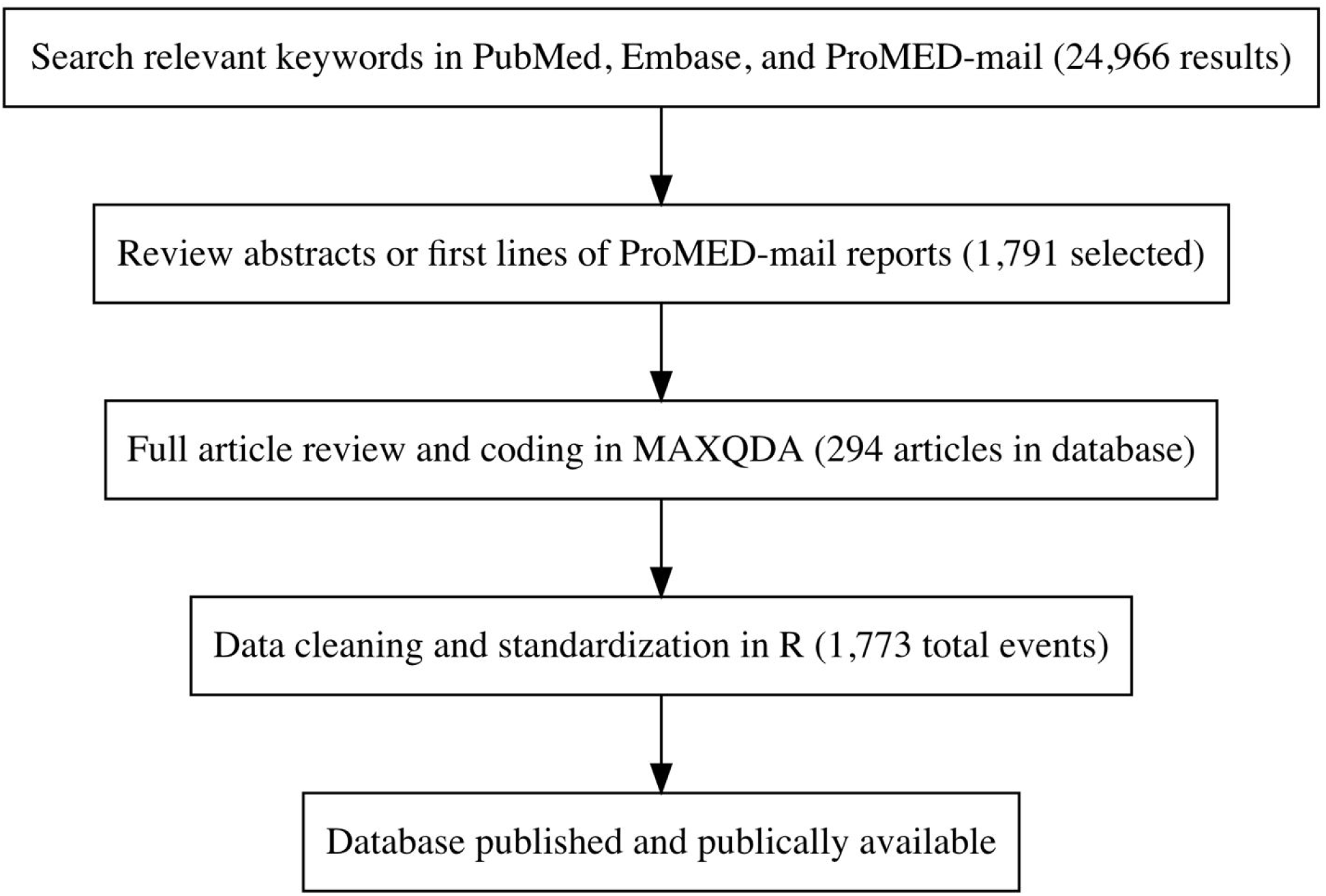
Overview of database building process

### Literature search

We drew from PubMed and Embase scientific literature databases and ProMED-mail to develop our database of events. PubMed and Embase collectively encompass a large fraction of English-language biomedical scientific literature, while ProMED-mail consists of clinical reports and news alerts that may not ultimately be published in the scientific press and have a faster reporting speed. We selected a recent ten-year period for scientific literature (2006-2017). ProMED-mail reports were drawn from a longer time span (1994-2017), as we found scientific articles published 2006-2017 frequently described events occurring many years before.

We searched for peer-reviewed manuscripts in PubMed and Embase published using the following terms: *‘antibiotic resistance’/exp OR (‘antibiotic’AND (‘resistant’ OR ‘resistance’)) OR ‘antimicrobial resistance’/exp OR (‘antimicrobial’AND (‘resistant’ OR ‘resistance’)) AND (first OR novel OR new OR emerging OR emergent) AND (case OR patient) AND [humans]/lim AND [2006-2017]/py*

This search yielded 23,770 results.

Because ProMED-mail searches use partial word stem-based matching, we used only the search terms ‘*antibiotic resistance ‘* and *‘antimicrobial resistance* ‘, which yielded 1,196 results. A total of 24,966 articles were compiled for screening.

### Abstract and Report Screening

We used the following inclusion criteria to screen the results of our literature search: the article must have described at least one **clinical case** (a) of infectious disease in a **patient** (b) caused by a novel (c) resistance (d) to a particular **antimicrobial drug or drug combination** (e) in **bacteria** (f). In this definition:

a. A “clinical case” is an individual who presents with symptoms to a medical professional and is determined by a medical professional to have been infected with the bacterium in question. Antimicrobial resistance in an asymptomatic individual’s commensal bacteria (as determined by a screening study, a “challenge” study, or a laboratory trial) does not meet the requirements of a clinical case.
b. The “patient” from which the bacterium of interest is identified must be human and may be of any age or gender. More than one patient can be included in an emergence event if all patients fell ill and were confirmed to be infected with a resistant bacterium at the same time, for example in the early stages of an outbreak.
c. “Novel” indicates that resistance to a given antimicrobial treatment in the bacterial species in question has not previously been detected or described in the country in question. In this definition, new mechanisms of resistance of a given bacteria to a given antimicrobial combination do not count as novel resistance (e.g. a novel plasmid carrying beta-lactamase in a bacterial species with previously described beta-lactam resistance.)
d. “Resistance” is the ability of the bacteria in question to survive standard antibacterial treatment against it. Survival is measured by the bacteria’s ability to continue to cause disease in its host or to spread to others for longer than the standard period following treatment. Standard treatments are determined by the WHO and by the countries in which the articles in the systematic review take place.
e. An “antimicrobial drug or drug combination” is a drug, drug class, or specific combination of drugs used to treat bacterial infections in humans. We focused exclusively on antibiotic resistance rather than resistance to other antimicrobials due to better data availability and based on the hypothesis that drivers for other antimicrobials might be different than for antibiotics.
f. “Bacteria” is a species or strain of pathogen in the domain Bacteria. This study does not assess the impact of resistance in viruses, fungi, or other pathogen types.

For results from the PubMed and Embase search, we programmatically downloaded abstracts and metadata. Article abstracts were manually reviewed by a team of screeners (all authors, see Author Contributions, below) to determine whether they likely contained a report of a novel emergence event, according to the above criteria. To ensure uniformity in abstract evaluation, all reviewers received training on the inclusion criteria and were required to achieve 90% agreement with a practice set of 100 previously-screened abstracts. Abstracts were screened separately by two individuals. Reviewers classified articles as “yes”, “maybe”, or “no” for inclusion. If both reviewers classified an article as “yes” or “maybe”, the article was downloaded as full-text for further review and to be coded for the database. The inter-scorer agreement rate was 82%. In cases when an article was marked as “no” by one of the reviewers, and “yes” by the other reviewer, a third reviewer was assigned to determine if the article should be included. 1,583 articles from PubMed and Embase passed review criteria and were downloaded for further review. The R package metagear^9^ was used to screen articles and manage screening data.

Full texts were downloaded for ProMED-mail search results, as ProMED-mail reports do not have abstracts. The first lines of each report were screened separately by two reviewers. If either reviewer classified a text as “yes” or “maybe”, it was selected for further review and to be coded for the database. 208 ProMED-mail texts passed this screening, contributing to a total of 1,791 total articles from PubMed, Embase, and ProMED-mail selected for review. Further details on the reproducible screening workflow are available in the data repository (see Data Records).

### Article coding

Articles from PubMed, Embase and ProMED-mail that passed screening were selected for full-text analysis, each by one reviewer, unless quality assurance checks required follow-up review (see Technical Validation). Reviewers read full-text articles to determine whether they fully met the case criteria, above, and to extract data by coding the text. Articles were excluded at this stage if it was found that they referred to or were duplicate reports of a previous emergence of the same drug-bacteria resistance within the country, if they reported on a non-bacterial pathogen, or if they did not identify the drug or bacterial species. A total of 294 articles were retained after full-text screening.

Articles were coded for four required fields: study country, drug name, bacteria, and event date. Drugs were coded to the lowest available taxonomic rank (i.e., specific drug rather than drug class, when available). Similarly, bacteria were coded at the species level when available. Where articles did not include an emergence location or date, location was inferred from the location of study authors’ institutions (often hospitals), and publication year was used for event date.

In addition to the required fields, articles were coded for the following secondary fields when available: patient attributes (age, gender, country of residence, recent travel locations, symptoms, comorbidities, and outcome), bacterial strains and markers, drug minimum inhibitory concentration (MIC) values, and hospital location (city, state/province). Screeners used MAXQDA^10^ software for article coding.

### Data Cleaning

We matched free-form values coded in article text to standardized values and ontologies. All locations were matched to Google Place names and geocoded. Country names were maintained according to article reporting and were not standardized to official country recognitions (e.g., United Nations member states). Drug names and bacteria were matched and standardized against the Medical Subject Headings (MeSH)^11^ and National Center for Biotechnology Information (NCBI) Organismal Classification^12^ ontologies, respectively, as provided by the Bioportal platform^13^. Where reported names did not exactly match ontologies or had ambiguous matches, we manually reviewed and corrected names to match the ontologies, in some cases requiring review of the original study to confirm accuracy. Dates were converted to ISO 8601 format. In cases when studies reported a range of dates, only the start date was included in the database. Other, optional fields (patient demographics, etc.) were not standardized in the current database release.

Data cleaning and standardization was performed in R version 3.6.1^14^, using the tidyverse framework^15^ for data manipulation. Geocoding used the Google Geocoding API via the ggmap R package^16^. Study dates were standardized using the lubridate R package^17^.

## Data Records

We present a database of 1,773 records of first clinical reports of unique bacterial-drug AMR detections by country, ranging from 1998 to 2017, drawn from 294 peer-reviewed articles and reports. While the ProMED-mail search extended to 1994, the first reported event that met our study criteria occurred in 1998. The database is available at DOI 10.5281/zenodo.3964895. In addition to the database, we present all intermediate data, including: article abstracts and full-text; raw exports from article coding; and a pre-filtered, pre-transformed database that contains all fields (primary and secondary) and events (including some non-emergence reports).

The database, ‘data-processed/events-db.csv’, contains the following fields:

- ‘ study_id’ - unique study identification number that can be joined with ‘ articles_db’ for study metadata.
- ‘ study_country’ - name of country where event occurred. Note that there are some studies that report on events in multiple countries.
- study_iso3c’ - three letter International Organization for Standardization (ISO) code
- ‘ study_location’ - full study location (including hospital, city, and state if available)
- ‘study_location_basis’ - spatial basis of study location (e.g., "hospital, city, state_province_district, country")
- ‘ residence_location’ - location of patient residence
- ‘travel_location’ - patient travel locations, if any reported. Multiple locations are separated by ‘;’.
- drug’ - antimicrobial drug, standardized to the Medical Subject Headings (MeSH) ontology^11^. Drug combinations are concatenated by ‘+’.
- ‘drug_rank’ - taxonomic classification of drug (i.e., drug name or group)
- ‘ segment_drug_combo’ - TRUE/FALSE resistance is to a combination of drugs
- drug_parent_name’ - name of the taxonomic parent of antimicrobial drug, standardized to the Medical Subject Headings (MeSH) ontology^11^.
- ‘bacteria’ - name of resistant bacteria, standardized to NCBI Organismal Classification ontology^12^.
- ‘bacteria_rank’ - taxonomic classification of bacteria name (e.g., “species”, “genus”)
- ‘bacteria_parent_name’ - name of the taxonomic parent of bacteria, standardized to NCBI Organismal Classification ontology^12^
- ‘bacteria_parent_rank’ - - taxonomic classification of bacteria parent name (e.g., “species”, “genus”)
- ‘ start-date’ - date that emergence was reported in format of *yyyy-mm-dd*
- ‘end_date’ - date that emergence was resolved, if reported, in format of *yyyy-mm-dd*
- ‘start_date_rank’ - specificity of the start date (i.e., year, month, day)

The file ‘data-processed/articles-db.csv’ contains metadata about each article, including citation information and full abstracts. This file can be joined with ‘data-processed/events-db.csv’ by the ‘study_id’ field. Coded full-text articles are in the ‘data-raw/coded-text-mex’ folder, and raw exports of these files in .xlsx form are in ‘data-raw/coded-segments’. The pre-filtered, pre-transformed database that contains all fields and events (including non-emergent events) is in ‘segments.csv’. All abstracts and ProMED-mail reports that were reviewed are in the ‘screening’ directory. Further details are provided in the data repository documentation.

We identified AMR emergence events in 66 countries, with the most reported events in the United States (152), India (129), and China (128) (**Figure 2**). Events were reported from 1998 through 2017, with the most events occurring in 2011 (**Figure 3**). See Usage Notes for discussion on the effects of reporting bias on the spatial and temporal coverage of the database.

**Figure 2.**
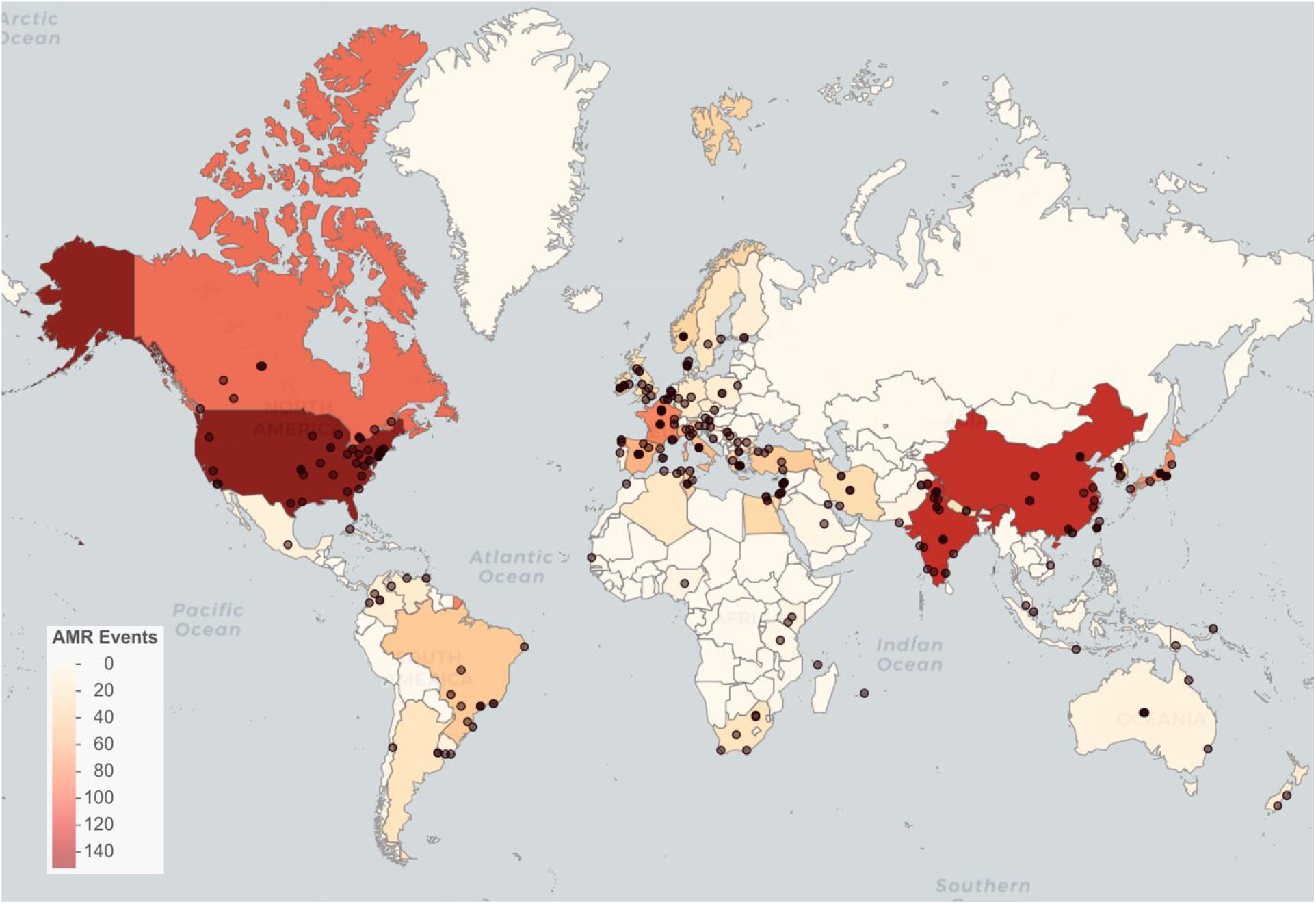
Global AMR emergence events. Points represent locations. Countries are shaded by event

**Figure 3.**
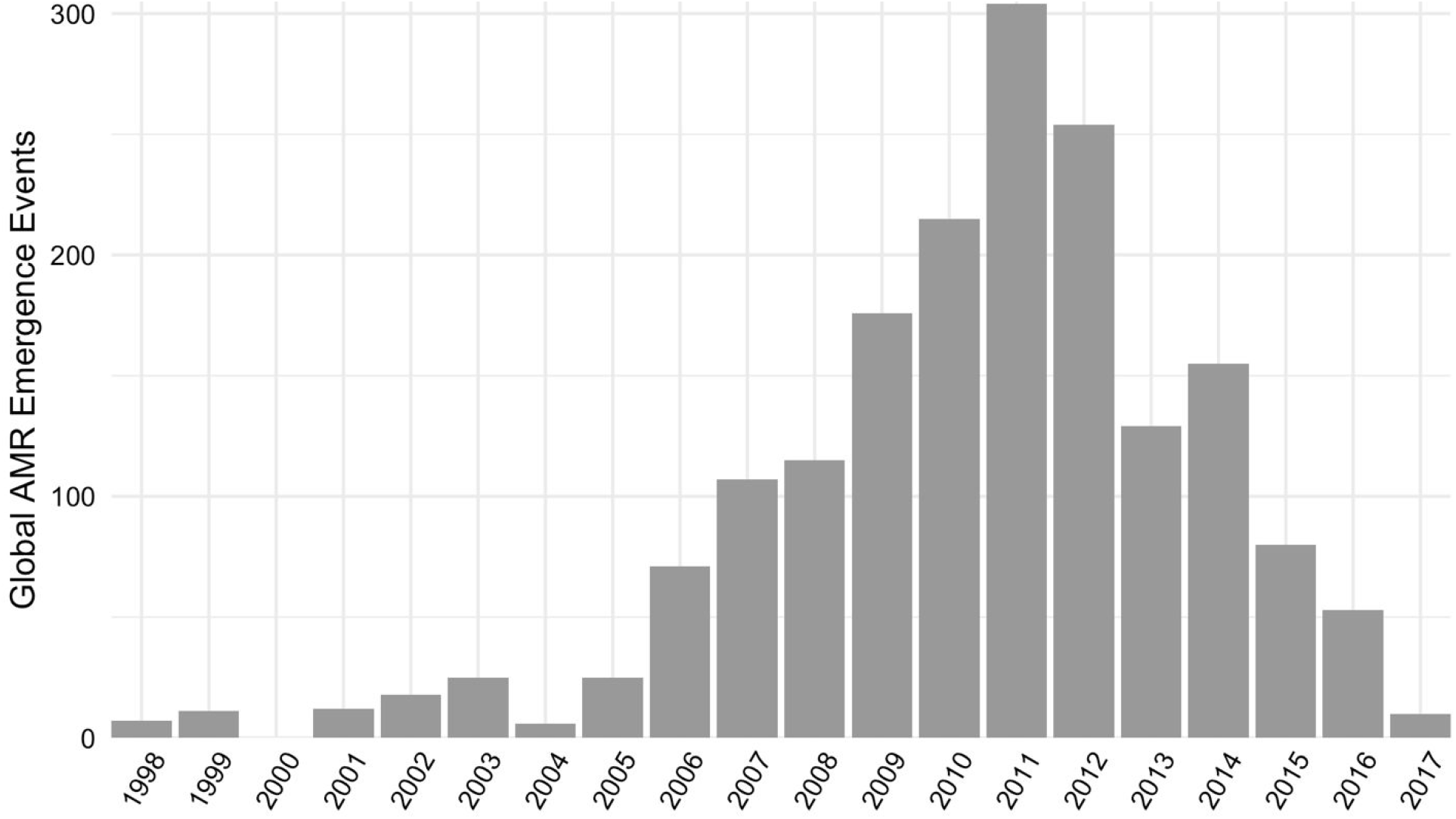
Count of global AMR emergence events in the database by year.

We found that *Klebsiella pneumoniae* and *Escherichia coli* were the most common bacteria in emergence events (**Figure 4**), supporting results from other databases that have found high rates of AMR prevalence for these species^3,8^. Of concern, our database indicates that both bacteria species had the greatest number of reports of novel resistance to imipenem and meropenem (**Figure 4**), which are carbapenem antibiotics often considered last resort treatment options for infections acquired in health care settings^3,18,19^. Also concerning were 23 cases of emergent resistance to colistin in 23 countries and 14 distinct bacteria, including *K. pneumoniae* and *E. coli*. Colistin is critically important for treating infections when no other options are available^19,20^.

**Figure 4.**
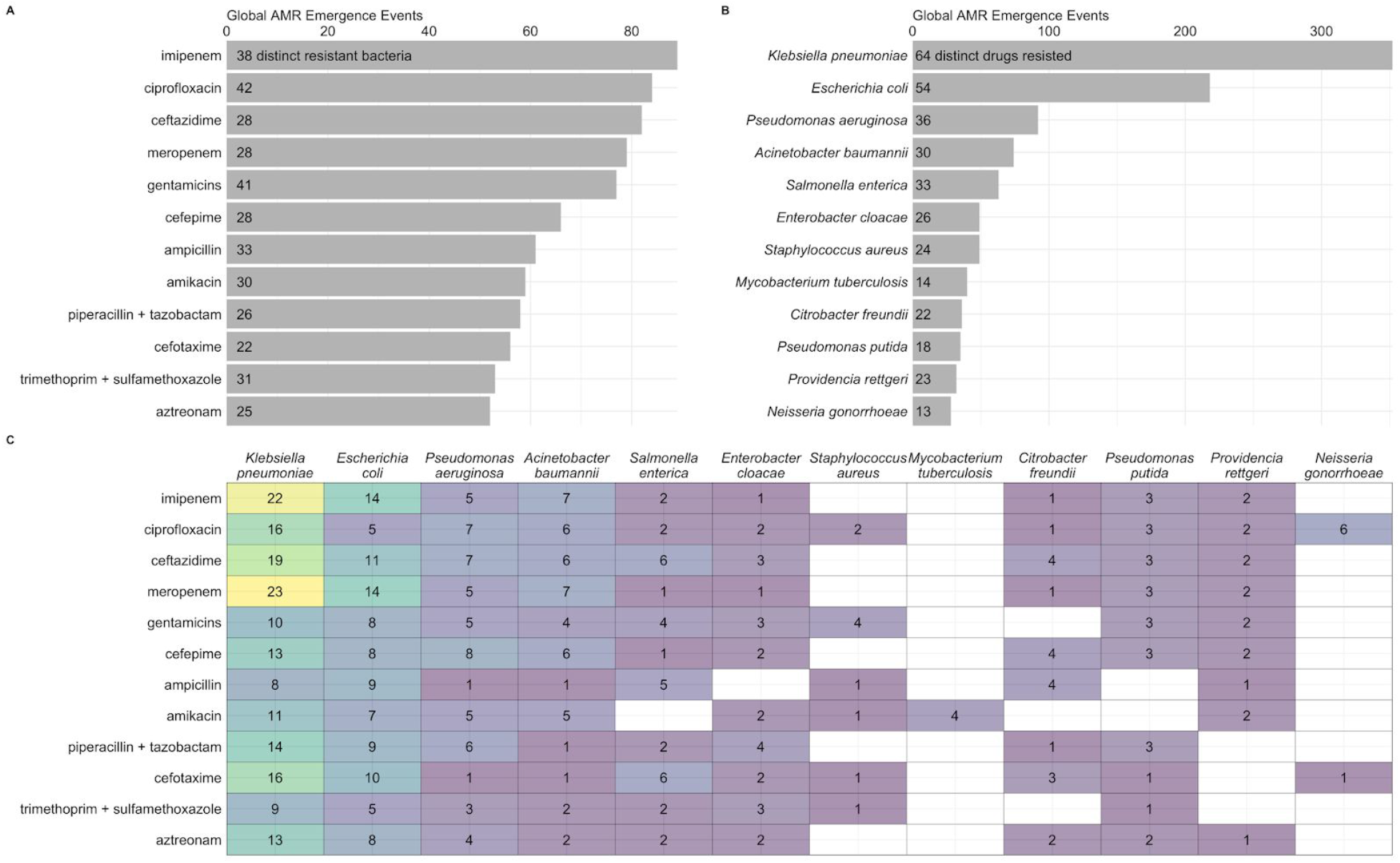
Global AMR emergence events for top twelve drugs (a) and bacteria (b), and for their combinations (c). Counts within bars represent distinct number of resistant bacteria (a) and drugs (b).

## Technical Validation

We implemented quality assurance checks throughout the data processing pipeline. We checked for errors in the MAXQDA article coding by confirming that all values were labeled and that links between values were properly assigned (e.g., links between drug names and MIC values). We checked for any studies missing study location, study date, drug, or bacteria, and manually revisited these articles to confirm missing fields. We also investigated any study reporting more than one location and/or date to confirm whether the study described multiple emergence events.

## Usage Notes

The database is developed using only English-language scientific literature and medical reporting events. It therefore represents events reported in this limited subset rather than truly representative clinical cases and strongly reflects reporting effort and practices. It is likely that AMR emergence events are systematically missing from the database for countries that are not English speaking and/or have less health care monitoring and reporting capacity, including those with lower GDP. Therefore, it is imperative that any analysis of the data account for the effects of reporting bias^21^.

Temporal patterns of AMR emergence events in the database reflect potentially incomplete coverage prior to 2006, as only ProMED-mail (not PubMed or Embase) was searched for this period. Further, the decrease in emergence events from 2011-2017 reflects lags between event occurrence and reporting.

There are several sources of ambiguity in the dataset due to imprecise reporting in literature (**Table 1**). While most studies reported the exact city or hospital of the emergence event (n = 200), others reported only state/province (n = 21) or study country (n = 73). Differences in geographic scales would need to be considered if using this dataset for spatial analysis. In addition, there are inconsistencies in studies’ reporting of drug and bacteria names. While most studies reported specific drug names (n = 87), others reported broad classes of drugs (n = 35). Study authors that reported classes of drugs may have administered multiple drugs to the patient(s), but because the specific drugs were not reported, we were only able to count the drug as part of a single event. Similarly, bacteria were primarily reported to the species level (n = 106), but some were reported only to the genus or family level (n = 5). In these cases, it is possible that the species was novel or not identifiable. Many studies that reported bacterial species also reported strains, and it is possible that there are differences in resistance by strain. However, due to inconsistent reporting, we have not standardized reported strains.

**Table 1.** Classification specificity of four primary database fields

|  | Classification | Count |
| --- | --- | --- |
| **Drug** | drug name | 84 |
|  | drug group | 35 |
|  | drug group/name combo | 3 |
| **Bacteria** | species | 106 |
|  | genus | 4 |
|  | family | 1 |
| **Location** | hospital | 123 |
|  | city | 77 |
|  | state/province/district | 21 |
|  | country | 73 |
| **Date** | day | 47 |
|  | month | 122 |
|  | year | 134 |

Finally, the database presents first AMR emergence events by country. Emergence events may be due to either mutation events or transport from other geographies. Analyses examining determinants of first emergence at a country level should consider import as a possible mechanism. Some studies reported locations of residence and recent travel of patients, and where available we have included this information, which may support such analysis. Analyses only examining first global emergence events should remove records of all but the earliest dates of each unique bacterial-drug combination.

This database, serving as a complement to existing databases that track resistance and spread, will allow researchers to target efforts for surveillance and interventions and to analyze factors that contribute to AMR emergence.

## Code availability

All code used to develop, code, and clean the database is available for download at https://github.com/ecohealthalliance/amr-db and archived on the Zenodo data repository [https://zenodo.org/record/3964895#.XzQEw2iYpD8].

## Data Availability

All data is publicly available

https://zenodo.org/record/3964895

## Acknowledgements

We thank Ayomide Sokale for screening abstracts for the database. This work was supported by the United States Agency for International Development (USAID) Emerging Pandemic Threats PREDICT project.

## Author Contributions

A. W., C.Z-T., and P.D. conceived the study. E.M., N.R., A.W., K.W., B.W.M., C.B., and E.J. contributed to study design and management. E.M., C.B., and K.W. wrote code to support project workflow and data cleaning. A.W., B.W.M., C.B., E.J., K.W., M.K., J.K., M.D., Z.M., and N.E. screened abstracts. A.W., B. W.M., E.M., M.D., S.D., S.M., and Z.M. coded articles and participated in manual data cleaning. E.M. and N.R. drafted the manuscript. A.W., B.W.M., E.J., and K.W., contributed text to the methods section. All authors were involved in editing and approving the manuscript.

## Competing Interests

The authors declare no competing interests.

## Notes

### Competing Interest Statement

The authors have declared no competing interest.

### Funding Statement

This work was made possible by the generous support of the American people through the United States Agency for International Development (USAID) Emerging Pandemic Threats PREDICT (Cooperative Agreement No. AID-OAA0A-14-00102). The contents are the responsibility of the authors and do not necessarily reflect the views or the policy of USAID or the United States Government.

